# Ischemic Stroke after Bivalent COVID-19 Vaccination: A Self-Controlled Case Series Study

**DOI:** 10.1101/2023.10.12.23296968

**Authors:** Stanley Xu, Lina S. Sy, Vennis Hong, Kimberly J. Holmquist, Lei Qian, Paddy Farrington, Katia J. Bruxvoort, Nicola P. Klein, Bruce Fireman, Bing Han, Bruno J. Lewin

**Author notes:** Corresponding author: Stanley Xu, PhD Department of Research & Evaluation Kaiser Permanente Southern California 100 S Los Robles Pasadena, California 91101.

## Abstract

**Introduction:** The potential association between bivalent COVID-19 vaccination and ischemic stroke remains uncertain, despite several studies conducted thus far. The purpose is to evaluate the risk of ischemic stroke following bivalent COVID-19 vaccination.

**Methods:** A self-controlled case series study was conducted among members aged ≥12 years who experienced ischemic stroke between September 1, 2022 and March 31, 2023 in a large California health care system. Ischemic strokes were identified using ICD-10 codes in Emergency Department and inpatient settings. Exposures were Pfizer-BioNTech or Moderna bivalent COVID-19 vaccination. Risk intervals were pre-specified as 1–21 days and 1–42 days after bivalent COVID-19 vaccination; all non-risk-interval person-time served as control interval. We conducted overall and subgroup analyses by age, history of SARS-CoV-2 infection, and co-administration of influenza vaccine. When an elevated risk was detected, we performed chart review of ischemic strokes, and re-evaluated the risk.

**RESULTS:** With 4933 cases, we found no increased risk within 21-day risk interval across vaccines and by subgroups. However, an elevated risk emerged within 42-day risk interval among individuals <65 years who received co-administration of Pfizer-BioNTech bivalent vaccine and influenza vaccine on the same day; relative incidence (RI) was 2.14 (95% CI, 1.02–4.49). Among those who also had history of SARS-CoV-2 infection, RI was 3.94 (95% CI, 1.10–14.16). After chart review, RIs were 2.35 (95% CI, 0.98–5.65) and 4.33 (95% CI, 0.98–19.11), respectively. Among individuals <65 years who received Moderna bivalent vaccine and had history of SARS-CoV-2 infection, RI was 2.62 (95% CI, 1.13–6.03) before chart review and 2.24 (95% CI, 0.78–6.47) after chart review.

**CONCLUSIONS:** The potential association between bivalent COVID-19 vaccination and ischemic stroke in the 1-42-day analysis warrants further investigation among individuals <65 years with influenza vaccine co-administration and prior SARS-CoV-2 infection.

## Introduction

On August 31, 2022, the U.S. Food and Drug Administration (FDA) granted emergency use authorizations (EUAs) for the Pfizer-BioNTech bivalent COVID-19 vaccine for individuals aged 12 years and older and the Moderna bivalent COVID-19 vaccine for individuals aged 18 years and older.^1,2^ The bivalent vaccines contain mRNA components derived from both the original strain of SARS-CoV-2 and the omicron variant BA.4 and BA.5 sublineages. Designed to be administered as a single booster dose, bivalent COVID-19 vaccines were recommended to be given ≥60 days after either primary vaccination or a monovalent booster dose.^2^ In the context of waning protection from primary vaccination, bivalent vaccines enhanced the immune response and boosted protection against the virus, offering an additional layer of defense for previously-vaccinated individuals.^3–5^

Safety data for bivalent mRNA COVID-19 vaccines were initially limited. Because the chemical components and production processes between monovalent and bivalent vaccines were similar, FDA granted EUAs for the bivalent COVID-19 vaccines based on safety data for monovalent vaccines as well as limited bivalent safety data from clinical trials.^1,2^ To monitor safety post-licensure, a study utilizing v-safe and the Vaccine Adverse Event Reporting System (VAERS) examined bivalent booster vaccinations in individuals aged ≥12 years and found that the safety profile was similar to that described for monovalent booster vaccinations.^6^ A recent study which comprehensively assessed potential adverse events associated with bivalent vaccines using TreeScan in the Vaccine Safety Datalink (VSD) network found no increased risk for a broad range of adverse events.^7^

The VSD has monitored COVID-19 vaccine safety since vaccinations began in December 2020.^8^ In late 2022, VSD’s rapid cycle analyses (RCA) detected a safety signal for ischemic stroke following the Pfizer-BioNTech COVID-19 bivalent booster vaccination among those 65 years and older, particularly among those who had received a bivalent booster dose and a high-dose or adjuvanted influenza vaccine on the same day (co-administration).^9^ The CDC and FDA announced this safety signal in January 2023.^10^ This safety signal attenuated as data accumulated.^11^ Another cohort study among adults aged 65 and older reported that those who received the Pfizer-BioNTech bivalent booster had a similar hazard for ischemic stroke encounters compared to those who received the Moderna bivalent booster vaccine, but had a lower hazard than those who received the Pfizer-BioNTech/Moderna monovalent boosters.^12^ In another study, compared to monovalent vaccination, bivalent vaccination was not found to be associated with increased risk of ischemic stroke, hemorrhagic stroke, myocardial infarction, and pulmonary embolism.^13^

The objective of this study was to assess the risk of ischemic stroke after bivalent COVID-19 vaccination among individuals enrolled in Kaiser Permanente Southern California (KPSC) using a modified self-controlled case series (SCCS) design. Subgroup analyses were also conducted by age (<65 years versus ≥65 years), history of SARS-CoV-2 infection, and co-administration of influenza vaccine.

## Methods

### Study Population and Study Period

We conducted a SCCS study among members aged ≥12 years from KPSC, a large integrated health care system in the US. The SCCS analytic datasets included individuals who experienced ischemic stroke events between September 1, 2022 and March 31, 2023, had completed a COVID-19 vaccine primary series, and had received their last monovalent dose ≥60 days before September 1, 2022. We required KPSC membership on September 1, 2022.

### Exposure and Observation Period

The exposure was defined as the administration of the Pfizer-BioNTech bivalent COVID-19 vaccine (for individuals aged ≥12 years) or the Moderna bivalent COVID-19 vaccine (for individuals aged ≥18 years) between September 1, 2022 and March 31, 2023. The observation period for the recipients of a bivalent COVID-19 vaccine started on September 1, 2022 and ended on March 31, 2023 or upon death, receipt of the second bivalent dose, or disenrollment, whichever came first.

To adjust for seasonality, we also included ischemic stroke events occurring among eligible individuals aged ≥12 years who had completed a primary series and had received their last monovalent dose ≥60 days before September 1, 2022, but who did not receive a bivalent vaccine during September 1, 2022 - March 31, 2023 (non-bivalent recipients [NBR]). The observation period for ischemic stroke events among NBR started on September 1, 2022, and ended on March 31, 2023, or upon death or disenrollment, whichever came first.

### Outcome

The outcome was defined as the first occurrence of an ischemic stroke event between September 1, 2022 and March 31, 2023.^14^ Ischemic stroke events were identified through medical encounters with an ICD-10 diagnosis code of G45.8, G45.9, or I63.* in the emergency department (ED) or inpatient settings. We also looked back 30 days prior to September 1, 2022, to ensure that the episode was incident. We excluded ischemic stroke events due to other possible causes and adjusted the onset date (details in eTable 1).

We considered these ischemic stroke events that were identified with ICD-10 codes to be electronically identified ischemic stroke events.

### Covariates

We collected demographic variables (age, sex, race/ethnicity) to describe the characteristics of the study population, as well as concomitant influenza vaccination during the study period and history of SARS-CoV-2 infection in the year prior to September 1, 2022.

### Statistical Analyses

We assessed the risk of ischemic stroke following the administration of the Pfizer-BioNTech and Moderna bivalent COVID-19 vaccines separately. Demographic characteristics of individuals who experienced ischemic stroke events during the study period were described among Pfizer-BioNTech bivalent vaccine recipients, Moderna bivalent vaccine recipients, and NBR.

The risk intervals were pre-specified as 1–21 days and 1–42 days after administration of bivalent COVID-19 vaccines, with person-time outside of these risk intervals serving as the control interval. The risk intervals started on the day of vaccination (Day 1). Because individuals who had ischemic stroke events might be likely to postpone or avoid bivalent vaccination, we used a modified SCCS approach for event-dependent exposures.^17^ The modified SCCS used a pseudo-likelihood approach in the counterfactual framework to estimate relative incidence (RI) and 95% confidence intervals (CI) of events comparing the risk intervals to their corresponding control intervals by maximizing a Poisson pseudo-likelihood. In the SCCS analyses, ischemic stroke events occurring among eligible individuals who did not receive the bivalent vaccines were included to account for seasonality, by incorporating calendar month into the model.^14^ Given that age did not significantly vary during the relatively short observation period of 7 months, it was not adjusted as a time-varying covariate.

Additionally, we performed several subgroup analyses based on age (<65 versus ≥65 years), co-administration of bivalent COVID-19 vaccine with same-day influenza vaccine (yes/no), and history of SARS-CoV-2 infection (confirmed by a positive laboratory test or a COVID-19 diagnosis) within one year prior to September 1, 2022. When a safety signal (i.e., the lower bound of the 95% CI for RI exceeded 1.0) was detected in analyses of electronically identified ischemic stroke events, we conducted chart review among recipients of bivalent COVID-19 vaccines to confirm ischemic stroke events and identify onset date to determine whether confirmed ischemic stroke events fell in the risk or control interval; confirmation rates were then calculated (number of confirmed events divided by number of electronically identified events reviewed). We did not conduct chart review on ischemic stroke events among NBR due to the large number of events in this group and limited resources. In analyses of confirmed ischemic stroke events among recipients of bivalent COVID-19 vaccines, we introduced a randomized allocation of confirmed case status to the NBR group. This allocation was guided by the confirmation rates observed among recipients of bivalent COVID-19 vaccines, as outlined by Xu et al.^18^ Five simulated datasets were generated to replicate the allocation process. SCCS analyses were conducted on each dataset, and the resulting estimates were aggregated using Rubin’s rule,^19^ which accounts for both the variability within individual datasets and the variability across the multiple datasets.

Attributable risk (AR) was calculated using the approach described in Farrington et al.

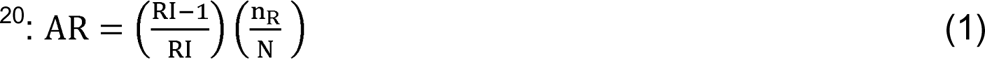

where RI is the relative incidence; n_R_ is the number of ischemic stroke events in the risk interval; and N is the number of recipients of a vaccine or dose. The reciprocal of AR is the number needed to harm (NNH).

Analyses were conducted using SAS version 9.4 (SAS Institute Inc., Cary, North Carolina) and the SCCS models were fitted with the R package SCCS.^21^ This study was approved by KPSC Institutional Review Board. Patients or members of the public were not involved in the design, or conduct, or reporting, or dissemination plans of the research.

## Results

Table 1 shows the characteristics of individuals who had ischemic stroke events. In total, there were 1057 ischemic stroke events among recipients of the Pfizer-BioNTech bivalent vaccine with a mean length of observation period of 204 days (ranging from 16 to 212 days), 827 ischemic stroke events among recipients of the Moderna bivalent vaccine with a mean length of observation period of 206 days (ranging from 31 to 212 days), and 3049 ischemic stroke events among NBR with a mean length of observation period of 197 days (ranging from 11 to 212 days). Notably, the majority of ischemic stroke events occurred among individuals ≥65 years.

**Table 1.**
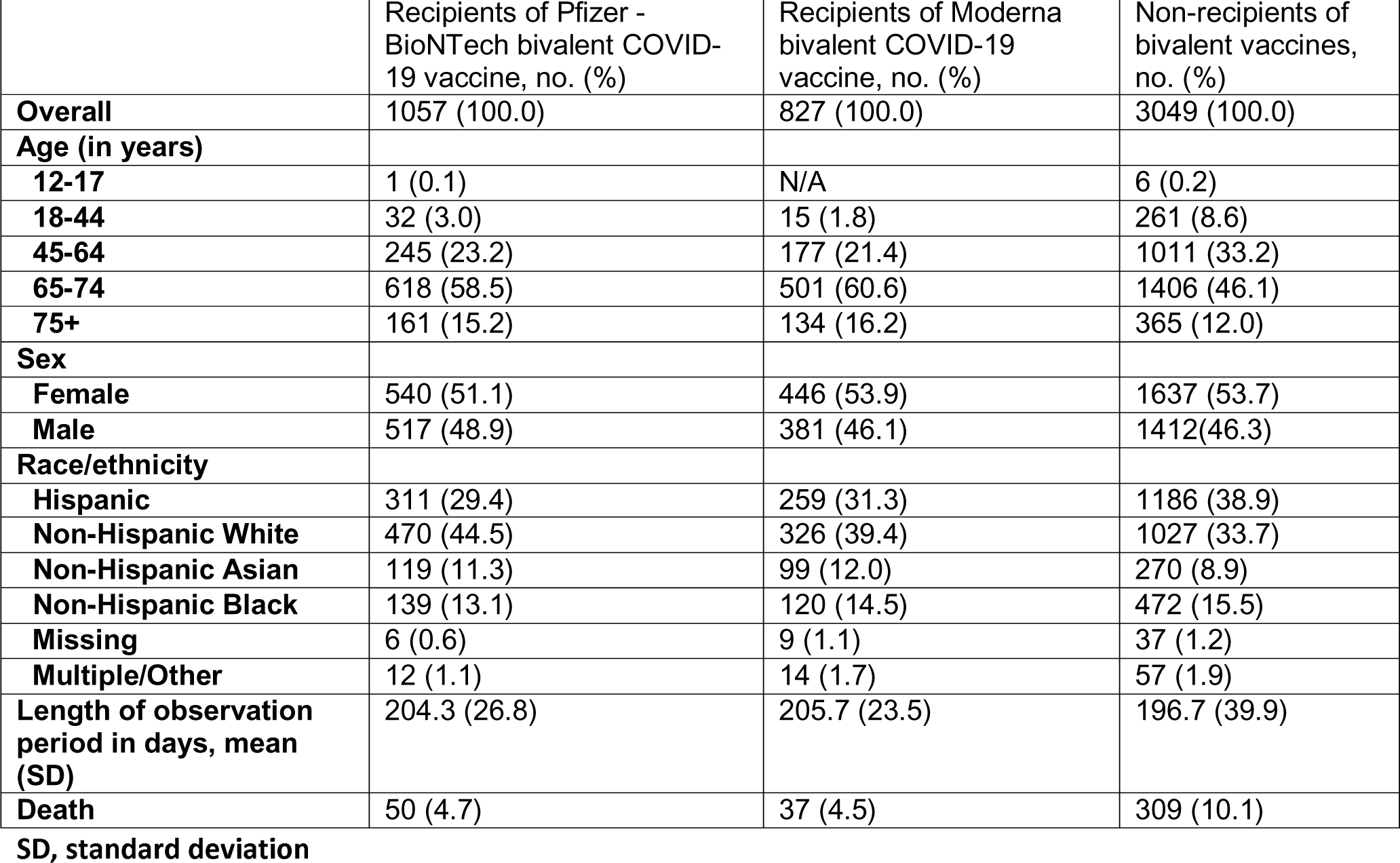
Characteristics of Individuals Who Had Ischemic Stroke Events during the Period from September 1, 2022 to March 31, 2023.

### Risk of Ischemic Stroke Following the Pfizer-BioNTech Bivalent COVID-19 Vaccine

For the Pfizer-BioNTech bivalent COVID-19 vaccine, there were 103 electronically identified ischemic stroke events in the 21-day post-vaccination risk interval, 954 events in the control interval, and 3049 events among NBR; the overall RI was 0.90 (95% CI, 0.73–1.12) (eTable 2). The RI was not significantly above 1 across all subgroup analyses by age, co-administration of influenza vaccine, and history of SARS-CoV-2 infection.

In analyses extending the risk interval to 1–42 days following bivalent vaccination, the overall RI was 0.97 (95% CI, 0.81–1.15) (Table 2). However, in subgroup analyses using the 1–42-day risk interval, we observed an increased risk of ischemic stroke only among individuals <65 years of age who also received an influenza vaccine on the same day. The RI in this subgroup was 2.14 (95% CI, 1.02–4.49). Among the subset who also had a documented history of SARS-CoV-2 infection within the year preceding the study period, the RI increased to 3.94 (95% CI, 1.10–14.16).

**Table 2.** Numbers of Electronically Identified Ischemic Stroke Events and Relative Incidences in the 42 Days after Pfizer-BioNTech Bivalent COVID-19 Vaccination.

|  | All ages |  |  |  | <65 years old |  |  |  | ≥65 years old |  |  |  |
| --- | --- | --- | --- | --- | --- | --- | --- | --- | --- | --- | --- | --- |
|  | Number of events |  |  |  | Number of events |  |  |  | Number of events |  |  |  |
|  | Risk interval | Control interval | NBR<sup>§</sup> | Relative incidence (95% CI) | Risk interval | Control interval | NBR<sup>§</sup> | Relative incidence (95% CI) | Risk interval | Control interval | NBR<sup>§</sup> | Relative incidence (95% CI) |
| **Overall** | 212 | 845 | 3049 | 0.97 (0.81–1.15) | 50 | 228 | 1278 | 0.98 (0.69–1.38) | 162 | 617 | 1771 | 0.96 (0.79–1.17) |
| With history of SARS-CoV-2<sup>□</sup> | 35 | 139 | 565 | 0.98 (0.66–1.47) | 15 | 53 | 292 | 1.22 (0.64–2.33) | 20 | 86 | 273 | 0.83 (0.50–1.40) |
| Without history of SARS-CoV-2 | 177 | 706 | 2484 | 0.97 (0.80–1.17) | 35 | 175 | 986 | 0.91 (0.61–1.38) | 142 | 531 | 1498 | 0.99 (0.80–1.22) |
| **Co-administration of influenza vaccine, overall** | 41 | 106 | 3049 | 0.99 (0.68–1.45) | 15 | 16 | 1278 | 2.14 (1.02–4.49) | 26 | 90 | 1771 | 0.77 (0.49–1.23) |
| With history of SARS-CoV-2<sup>□</sup> | 9 | 18 | 565 | 1.36 (0.61–3.05) | 6 | 4 | 292 | 3.94 (1.10–14.16) | 3 | 14 | 273 | 0.58 (0.16–2.14) |
| Without history of SARS-CoV-2 | 32 | 88 | 2484 | 0.92 (0.60–1.41) | 9 | 12 | 986 | 1.58 (0.61–4.09) | 23 | 76 | 1498 | 0.81 (0.49–1.33) |
| **No co-administration of influenza vaccine, overall** | 171 | 739 | 3049 | 0.95 (0.79–1.16) | 35 | 212 | 1278 | 0.80 (0.54–1.19) | 136 | 527 | 1771 | 1.00 (0.81–1.24) |
| With history of SARS-CoV-2<sup>□</sup> | 26 | 121 | 565 | 0.87 (0.55–1.38) | 9 | 49 | 292 | 0.82 (0.37–1.80) | 17 | 72 | 273 | 0.85 (0.49–1.50) |
| Without history of SARS-CoV-2 | 145 | 618 | 2484 | 0.98 (0.80–1.21) | 26 | 163 | 986 | 0.81 (0.51–1.28) | 119 | 455 | 1498 | 1.03 (0.81–1.30) |

<sup>§</sup>Non-bivalent recipients (NBR) were eligible individuals who did not receive a bivalent vaccine but had completed a primary series of COVID-19 vaccination and had their last monovalent dose ≥60 days before 9/1/2022. Inclusion of these events helps to adjust for temporal trends (seasonality). The same NBR population was used in overall bivalent analyses as well as bivalent analyses stratified by co-administration of influenza vaccine. <sup>□</sup> Had SARS-CoV-2 infection (i.e., SARS-CoV-2 positive laboratory test or a COVID-19 diagnosis) during the year prior (08/31/2021–08/31/2022).

In this 1–42-day risk interval analysis of the specific subgroup of individuals aged <65 years who received bivalent and influenza vaccines on the same day, chart review of the 31 electronically identified ischemic stroke events found that 2 were determined to be hemorrhagic strokes, and 8 were subsequently found to not meet the criteria for true ischemic stroke events, yielding a confirmation rate of 68%. With the verified 21 ischemic stroke events and ischemic stroke events among NBR (not verified through chart review, but adjusted for using a 68% confirmation rate), we proceeded to re-evaluate the RI in this subgroup. The number of confirmed ischemic stroke events was graphed over the interval in days between bivalent vaccination and ischemic stroke event (Figure 1). There were 10 events in the 1–42-day risk interval and 11 events in the control interval. Using a risk interval of 1–42 days after co-administration of the Pfizer-BioNTech bivalent vaccine and influenza vaccine, the overall RI derived from analyzing confirmed ischemic stroke events among those aged <65 years was 2.35 (95% CI, 0.98–5.65; *P*=.06) (Table 3). Between September 1, 2022 and March 31, 2023, 117,423 individuals aged <65 years received Pfizer-BioNTech bivalent vaccine and influenza vaccine on the same day. According to equation (1), AR = 4.89 x 10^-5^ and NNH = 20,440 with a risk interval of 1–42 days. Among 21,128 individuals who also had a documented history of SARS-CoV-2 infection within the year preceding the study period, the RI increased to 4.33 (95% CI, 0.98–19.11; *P*=.05) (Table 3); according to equation 1, AR = 1.46 x 10^-4^ and NNH = 6,868 with a risk interval of 1–42 days. Among the 10 confirmed ischemic stroke events in the risk interval of 1–42 days, the mean age was 58 years, ranging from 48 to 63 years. Among these cases, 2 individuals had a documented history of previous ischemic stroke, and no one died as of March 31, 2023. In addition, 7 received the standard dose, egg-based quadrivalent influenza vaccine, while 3 received an influenza vaccine of unknown formulation.

**Figure 1.**
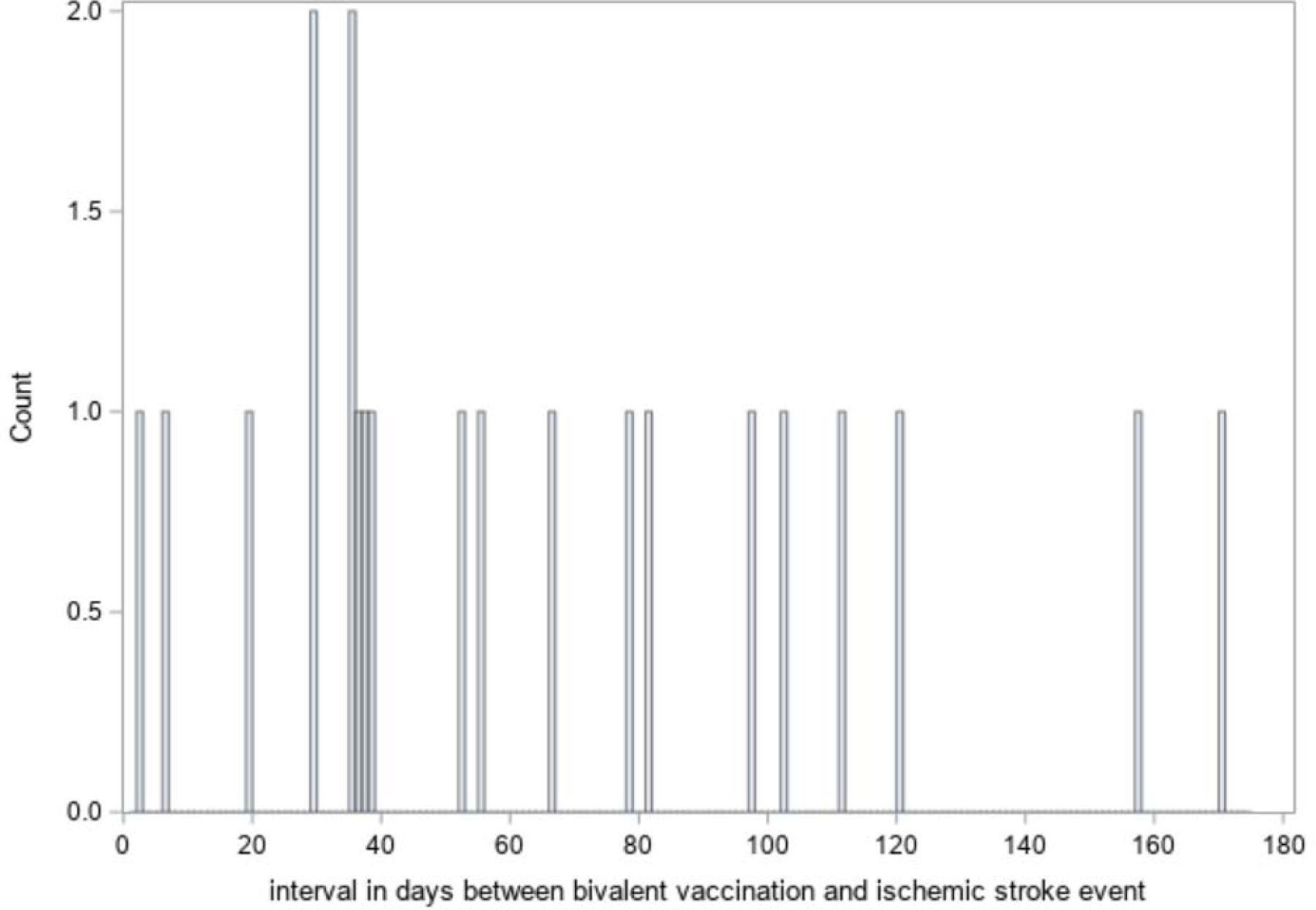
Number of Confirmed Ischemic Stroke Events over the Interval in Days between Bivalent Vaccination and Ischemic Stroke Event among Those Who Received Pfizer-BioNTech Bivalent Vaccine and Influenza Vaccine on the Same Day

**Table 3.** Numbers of Confirmed Ischemic Stroke Events among Recipients of the Pfizer-BioNTech Bivalent COVID-19 Vaccine Aged <65 years, and Relative Incidences in the 42 Days after Co-administration of Bivalent and Influenza Vaccines.

|  | Number of events |  |  |  |
| --- | --- | --- | --- | --- |
|  | Risk interval | Control interval | NBR<sup>§</sup> | Relative incidence (95% CI) |
| **Co-administration of influenza vaccine, overall** | 10 | 11 | 874 | 2.35 (0.98–5.65) |
| With history of SARS-CoV-2<sup>□</sup> | 4 | 3 | 197 | 4.33 (0.98–19.11) |
| Without history of SARS-CoV-2 | 6 | 8 | 677 | 1.76 (0.57–5.42) |

<sup>§</sup>Non-bivalent recipients (NBR) were eligible individuals who did not receive a bivalent vaccine but had completed a primary series of COVID-19 vaccination and had their last monovalent dose ≥60 days before 9/1/2022. Inclusion of these events helps to adjust for temporal trends (seasonality). A confirmation rate of 68% was applied to ischemic stroke events among NBR.
<sup>□</sup> Had SARS-CoV-2 infection (i.e., SARS-CoV-2 positive laboratory test or a COVID-19 diagnosis) during the year prior (08/31/2021–08/31/2022).

### Risk of Ischemic Stroke Following the Moderna Bivalent COVID-19 Vaccine

When using a risk interval of 21 days following Moderna bivalent vaccination, the overall risk of ischemic stroke was not elevated from the analysis of electronically identified ischemic stroke events (RI=0.91; 95% CI, 0.71–1.15), a finding that held true across all subgroup analyses by age, co-administration of influenza vaccine, and history of SARS-CoV-2 infection (eTable 3). However, extending the risk interval to 42 days after Moderna bivalent vaccination showed an increased risk of ischemic stroke among individuals <65 years of age who had a documented history of SARS-CoV-2 infection, with an RI of 2.62 (95% CI, 1.13–6.03). This subgroup involved a total of 36 ischemic stroke events among recipients of the Moderna bivalent COVID-19 vaccine (Table 4).

**Table 4.**
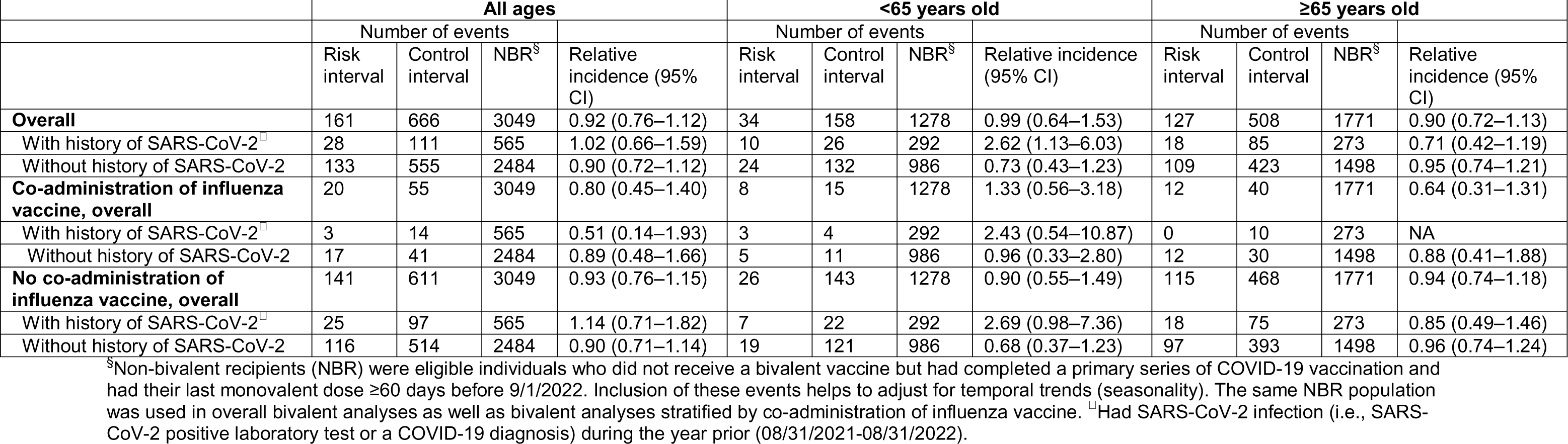
Numbers of Electronically Identified Ischemic Stroke Events and Relative Incidences in the 42 Days after Moderna Bivalent COVID-19 Vaccination.

|  | All ages |  |  |  | <65 years old |  |  |  | ≥65 years old |  |  |  |
| --- | --- | --- | --- | --- | --- | --- | --- | --- | --- | --- | --- | --- |
|  | Number of events |  |  |  | Number of events |  |  |  | Number of events |  |  |  |
|  | Risk interval | Control interval | NBR<sup>§</sup> | Relative incidence (95% CI) | Risk interval | Control interval | NBR<sup>§</sup> | Relative incidence (95% CI) | Risk interval | Control interval | NBR<sup>§</sup> | Relative incidence (95% CI) |
| **Overall** | 161 | 666 | 3049 | 0.92 (0.76–1.12) | 34 | 158 | 1278 | 0.99 (0.64–1.53) | 127 | 508 | 1771 | 0.90 (0.72–1.13) |
| With history of SARS-CoV-2<sup>□</sup> | 28 | 111 | 565 | 1.02 (0.66–1.59) | 10 | 26 | 292 | 2.62 (1.13–6.03) | 18 | 85 | 273 | 0.71 (0.42–1.19) |
| Without history of SARS-CoV-2 | 133 | 555 | 2484 | 0.90 (0.72–1.12) | 24 | 132 | 986 | 0.73 (0.43–1.23) | 109 | 423 | 1498 | 0.95 (0.74–1.21) |
| **Co-administration of influenza vaccine, overall** | 20 | 55 | 3049 | 0.80 (0.45–1.40) | 8 | 15 | 1278 | 1.33 (0.56–3.18) | 12 | 40 | 1771 | 0.64 (0.31–1.31) |
| With history of SARS-CoV-2<sup>□</sup> | 3 | 14 | 565 | 0.51 (0.14–1.93) | 3 | 4 | 292 | 2.43 (0.54–10.87) | 0 | 10 | 273 | NA |
| Without history of SARS-CoV-2 | 17 | 41 | 2484 | 0.89 (0.48–1.66) | 5 | 11 | 986 | 0.96 (0.33–2.80) | 12 | 30 | 1498 | 0.88 (0.41–1.88) |
| **No co-administration of influenza vaccine, overall** | 141 | 611 | 3049 | 0.93 (0.76–1.15) | 26 | 143 | 1278 | 0.90 (0.55–1.49) | 115 | 468 | 1771 | 0.94 (0.74–1.18) |
| With history of SARS-CoV-2<sup>□</sup> | 25 | 97 | 565 | 1.14 (0.71–1.82) | 7 | 22 | 292 | 2.69 (0.98–7.36) | 18 | 75 | 273 | 0.85 (0.49–1.46) |
| Without history of SARS-CoV-2 | 116 | 514 | 2484 | 0.90 (0.71–1.14) | 19 | 121 | 986 | 0.68 (0.37–1.23) | 97 | 393 | 1498 | 0.96 (0.74–1.24) |

<sup>§</sup>Non-bivalent recipients (NBR) were eligible individuals who did not receive a bivalent vaccine but had completed a primary series of COVID-19 vaccination and had their last monovalent dose ≥60 days before 9/1/2022. Inclusion of these events helps to adjust for temporal trends (seasonality). The same NBR population was used in overall bivalent analyses as well as bivalent analyses stratified by co-administration of influenza vaccine. <sup>□</sup>Had SARS-CoV-2 infection (i.e., SARS-CoV-2 positive laboratory test or a COVID-19 diagnosis) during the year prior (08/31/2021-08/31/2022).

Of the 36 ischemic stroke events, one was a hemorrhagic stroke and 12 were not confirmed as true events through medical chart review, yielding a confirmation rate of 64%. After using a risk interval of 1–42 days following the Moderna bivalent vaccination and applying a 64% confirmation rate to ischemic stroke events among NBR, the RI derived from analyzing confirmed ischemic stroke events among those aged <65 years who had a documented history of SARS-CoV-2 infection was 2.24 (95% CI, 0.78–6.47; *P*=.14).

## Discussion

These SCCS analyses did not find evidence that the risk of ischemic stroke was elevated during the 1–21-day post-vaccination risk interval in both overall and subgroup analyses by age (<65 years versus ≥65 years), prior history of SARS-CoV-2 infection, and co-administration of influenza vaccine, for both Pfizer-BioNTech and Moderna bivalent vaccines. However, based on electronically identified events, the risk of ischemic stroke was increased within the 1–42-day window after vaccination among those aged <65 years who received their Pfizer-BioNTech bivalent vaccine and influenza vaccine on the same day; the risk was even higher among those who also had a documented SARS-CoV-2 infection history. After conducting chart review of ischemic stroke events, the point estimate for the risk of ischemic stroke was still elevated in a risk interval of 1–42 days for these two subgroup analyses, but did not meet the threshold for statistical significance (*P*=.06 and .05, respectively).

For Moderna bivalent vaccination, an initial increase in the risk of ischemic stroke emerged within the 1–42-day window after vaccination among those aged <65 years who had a documented SARS-CoV-2 infection history. However, after conducting chart review of ischemic stroke events, the relative incidence was 2.24 but was no longer statistically significantly elevated possibly due to a decreased sample size (*P*=.14).

Our study showed an increased point estimate for the risk of ischemic stroke in a risk interval of 1–42 days only among those aged <65 years who received their Pfizer-BioNTech bivalent vaccine and influenza vaccine on the same day, although not statistically significant. This finding is unique and may be attributed to differences in the study design compared to previous studies. First, our study employed a calendar-based observation period spanning from September 1, 2022, to March 31, 2023. This extended timeframe enabled us to use a longer risk window of 1–42 days following vaccination in addition to the risk interval of 1–21 days in previous studies. Second, we did not exclude individuals with a history of ischemic stroke, but we did apply criteria to increase the likelihood that ischemic stroke events during the study period represented a new ischemic stroke episode. Nevertheless, it is possible that there was interaction between bivalent vaccination and history of ischemic stroke. Furthermore, in subgroup analyses, we considered the influence of history of SARS-CoV-2 infection. SARS-CoV-2 infection is associated with an increased risk of ischemic stroke,^15,16^ and risk factors for SARS-CoV-2 infection may overlap with risk factors for ischemic stroke. There is potential interaction between bivalent vaccination and history of SARS-CoV-2 infection. The finding that the point estimate for the risk of ischemic stroke was elevated among individuals aged <65 years but not among individuals aged ≥65 years is also biologically plausible. This may be due to the relatively heightened immune response and subsequent inflammation in the younger age group versus the older age group, and the fact that inflammation has been shown to be associated with an increased risk of ischemic stroke.^22,23^ Moreover, a smaller proportion of younger adults opted for bivalent vaccination,^24^ and those who did might have had a higher prevalence of comorbidities or poorer overall health status.

### Limitations and Strengths

This study had several limitations. First, the study took place in a single health care system and thus the sample size was smaller than that of the VSD COVID-19 vaccine RCA. Additionally, the number of ischemic stroke events in individuals with a documented SARS-CoV-2 infection history who received co-administration of Pfizer-BioNTech bivalent vaccine and influenza vaccine was very small. This raises concerns about the validity of the asymptotic large sample assumptions that underlie both the 95% confidence intervals and *P* values. Second, we did not conduct chart review of ischemic stroke events among NBR; these events contributed to establishing baseline rates of ischemic stroke events during the study period. In addressing this issue, when analyzing chart-confirmed ischemic stroke events among recipients of bivalent vaccine, we applied the confirmation rate of ischemic stroke events among recipients of bivalent vaccine to ischemic stroke events among NBR. Moreover, we also did not undertake chart review of ischemic stroke events from those analyses when safety signals were absent. Third, while we excluded ischemic stroke events occurring within 30 days of SARS-CoV-2 infection, it is possible that some ischemic stroke events included in the analyses involved individuals with asymptomatic or mild COVID-19 disease who did not have a documented SARS-CoV-2 infection. Fourth, the elevated point estimate for the risk of ischemic stroke, while not statistically significant, was observed within the 1–42-day period following co-administration of the Pfizer-BioNTech bivalent vaccine and influenza vaccine. This risk interval was longer than the 1–21 days or 1–28 days investigated in earlier research.^10,13,25^ However, the biological plausibility for the occurrence of a vaccine-related ischemic stroke beyond 28 days remains uncertain. Fifth, unaccounted time-varying confounders could have also influenced the findings. Finally, our analysis did not adjust for multiple subgroup analyses by age, co-administration of bivalent COVID-19 vaccine and influenza vaccine, and history of SARS-CoV-2 infection. These specific subgroup analyses were pre-specified due to their potential safety concerns. The decision not to make multiple comparison adjustments was deliberate, aimed at ensuring that any potential vaccine safety concern could be detected.

The study also had several strengths. First, we addressed the impact of previous ischemic stroke events on bivalent COVID-19 vaccination by employing an event-dependent modified SCCS design. Second, we explored effect heterogeneity by conducting subgroup analyses based on factors such as age, documented history of SARS-CoV-2 infection, and co-administration of influenza vaccine. Third, to adjust for temporal trends, we included ischemic stroke events among NBR. This strategy not only enhanced the accuracy of estimating the baseline rate but also improved the statistical power for identifying potential safety signals. Finally, we re-analyzed ischemic stroke events that were confirmed through chart review for those analyses where safety signals were detected.

Future research should include several key aspects to further enhance the validity and robustness of our findings. Collaborative efforts with additional health care systems will enable us to significantly increase our sample size. A larger sample size could provide sufficient statistical power to conduct sensitivity analyses such as exclusion of transient ischemic attack (TIA) and exclusion of those who had a history of ischemic stroke.

## Conclusions

We found no evidence to suggest that the Pfizer-BioNTech bivalent vaccine increased the risk of ischemic stroke among individuals aged >65 years, consistent with the attenuated signal from the VSD surveillance that motivated this study. We found an elevated point estimate for the risk of ischemic stroke within 1–42 days (but not within 1-21 days) after the co-administration of the Pfizer-BioNTech bivalent vaccine and influenza vaccine among individuals <65 years old that did not reach statistical significance, although the sample size was limited. Future studies with a larger sample size are needed to evaluate the association between bivalent COVID-19 vaccination and ischemic stroke, as well as contributing factors such as history of SARS-CoV-2 infection. Any potential risks of ischemic stroke associated with bivalent COVID-19 vaccination must be balanced against the potential benefits of bivalent COVID-19 vaccination in preventing COVID-19-associated ischemic stroke and severe COVID-19 disease.

## Disclaimer

The findings and conclusions in this report are those of the authors and do not necessarily represent the official views of the National Institutes of Health.

## Additional Contributions

Kaiser Permanente Southern California stroke chart abstractions from the Vaccine Safety Datalink were used in this work. Kristin Goddard and Pat Ross from Kaiser Permanente Northern California and Kayla Hanson from Marshfield Clinic Research Institute developed the stroke chart abstraction forms. We thank the KPSC chart abstractors Jose Pio and Julliane Bacerdo.

## Data Availability

All data produced in the present study are available upon reasonable request to the authors.

## SUPPLEMENT

**eTable 1.**
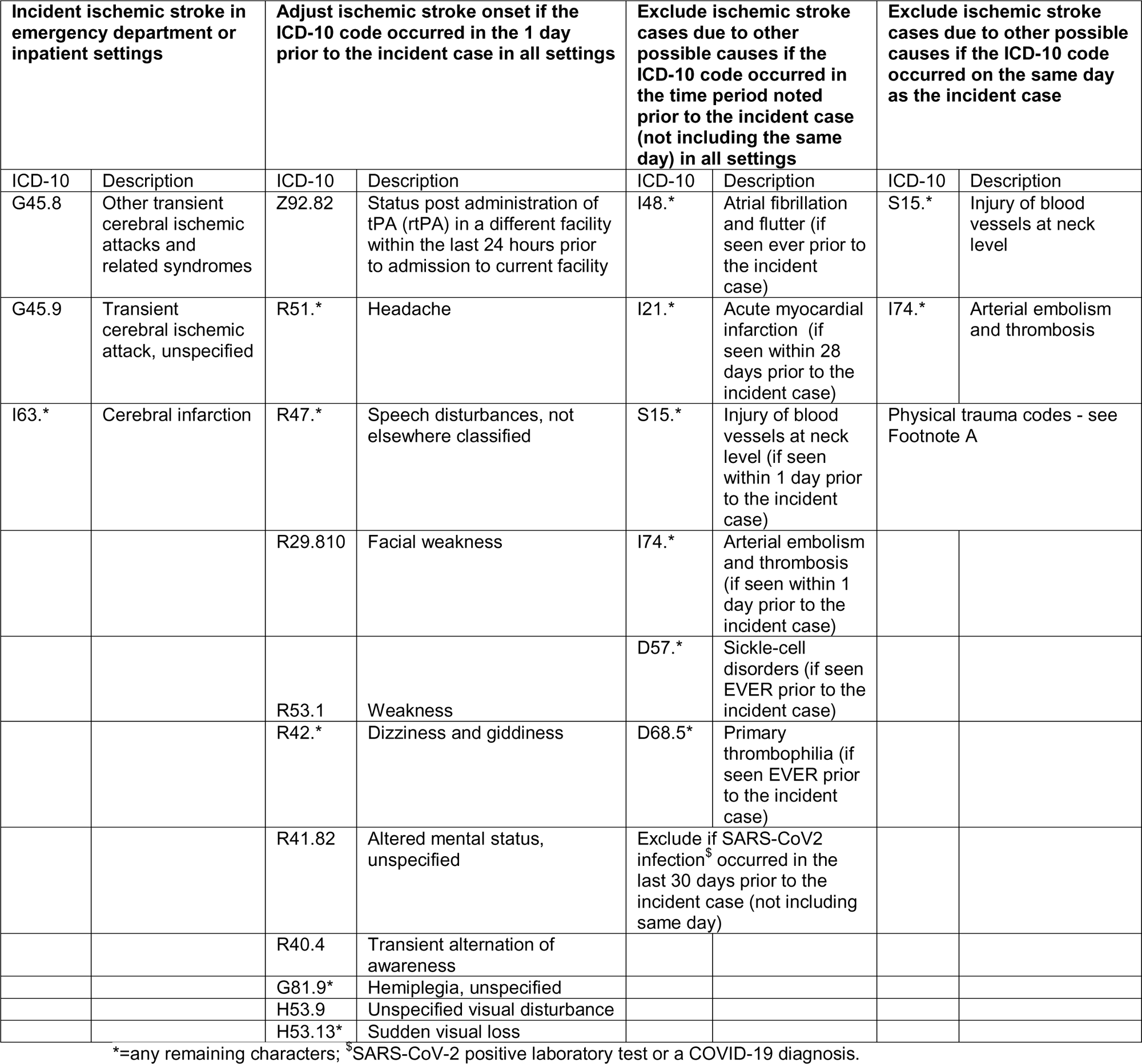
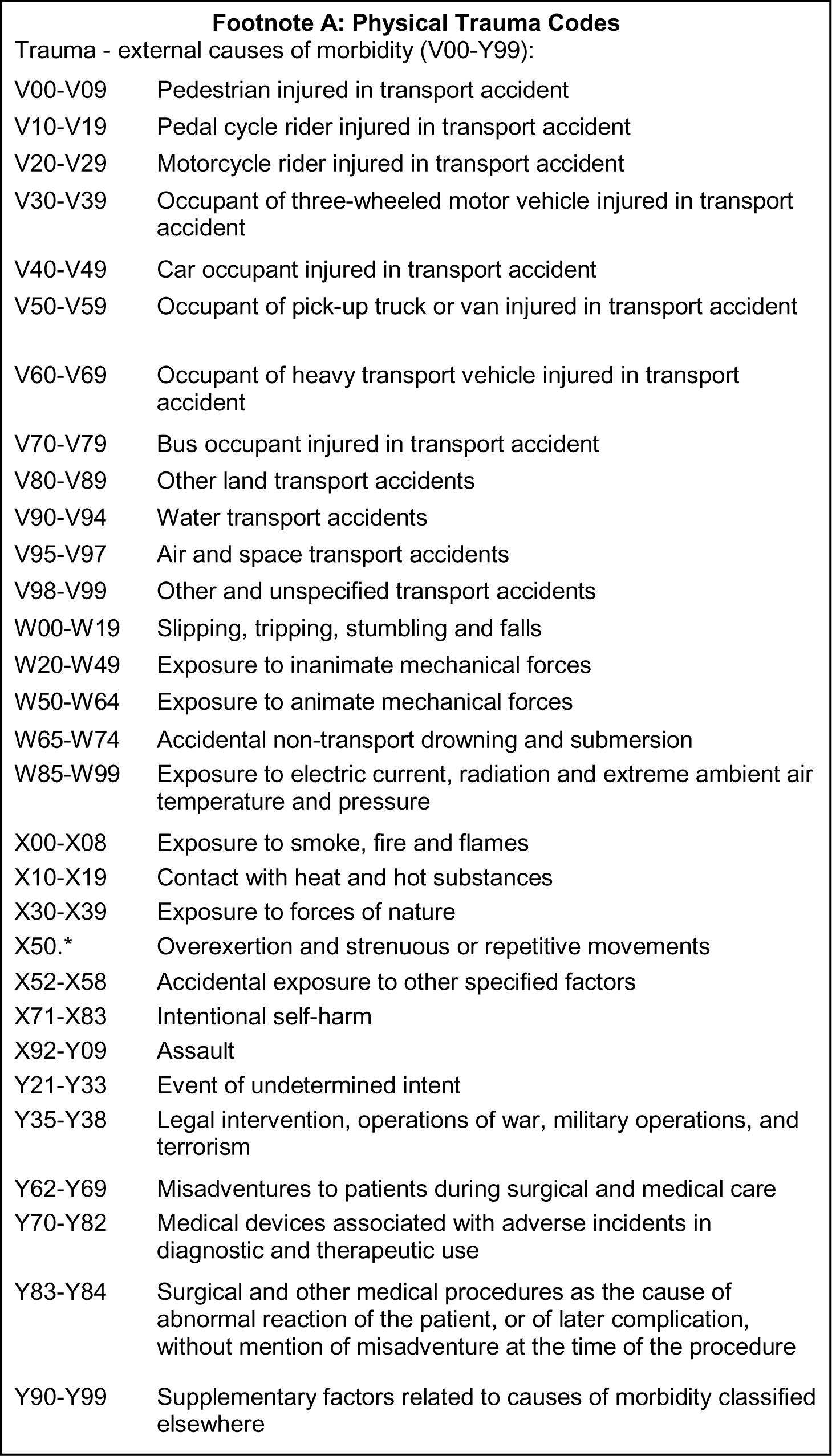
Definition of Electronically Identified Ischemic Strokes.

**eTable 2.** Numbers of Electronically Identified Ischemic Stroke Events and Relative Incidences in the 21 Days after Pfizer-BioNTech Bivalent COVID-19 Vaccination.

|  | All ages |  |  |  | <65 years old |  |  |  | ≥65 years old |  |  |  |
| --- | --- | --- | --- | --- | --- | --- | --- | --- | --- | --- | --- | --- |
|  | Number of events |  |  |  | Number of events |  |  |  | Number of events |  |  |  |
|  | Risk interval | Control interval | NBR<sup>§</sup> | Relative incidence (95% CI) | Risk interval | Control interval | NBR<sup>§</sup> | Relative incidence (95% CI) | Risk interval | Control interval | NBR<sup>§</sup> | Relative incidence (95% CI) |
| **Overall** | 103 | 954 | 3049 | 0.90 (0.73–1.12) | 19 | 259 | 1278 | 0.68 (0.42–1.12) | 84 | 695 | 1771 | 0.98 (0.77–1.25) |
| With history of SARS-CoV-2<sup>□</sup> | 12 | 162 | 565 | 0.68 (0.39–1.19) | 2 | 66 | 292 | 0.52 (0.18–1.51) | 10 | 96 | 273 | 0.77 (0.40–1.47) |
| Without history of SARS-CoV-2 | 91 | 792 | 2484 | 0.96 (0.76–1.21) | 17 | 193 | 986 | 0.75 (0.43–1.32) | 74 | 599 | 1498 | 1.02 (0.79–1.33) |
| **Co-administration of influenza vaccine, overall** | 22 | 125 | 3049 | 1.03 (0.65–1.62) | 6 | 25 | 1278 | 1.39 (0.60–3.24) | 16 | 100 | 1771 | 0.94 (0.55–1.62) |
| With history of SARS-CoV-2<sup>□</sup> | 3 | 24 | 565 | 0.85 (0.31–2.29) | 1 | 9 | 292 | 1.35 (0.33–5.52) | 2 | 15 | 273 | 0.61 (0.16–2.35) |
| Without history of SARS-CoV-2 | 19 | 101 | 2484 | 1.07 (0.65–1.79) | 5 | 16 | 986 | 1.41 (0.49–4.02) | 14 | 85 | 1498 | 1.02 (0.56–1.83) |
| **No co-administration of influenza vaccine, overall** | 81 | 829 | 3049 | 0.87 (0.68–1.11) | 13 | 234 | 1278 | 0.55 (0.30–1.01) | 68 | 595 | 1771 | 0.97 (0.75–1.27) |
| With history of SARS-CoV-2<sup>□</sup> | 9 | 138 | 565 | 0.63 (0.32–1.22) | 1 | 57 | 292 | 0.33 (0.07–1.50) | 8 | 81 | 273 | 0.78 (0.37–1.62) |
| Without history of SARS-CoV-2 | 72 | 691 | 2484 | 0.93 (0.71–1.20) | 12 | 177 | 986 | 0.64 (0.33–1.24) | 60 | 514 | 1498 | 1.01 (0.76–1.35) |

<sup>§</sup>Non-bivalent recipients (NBR) were eligible individuals who did not receive a bivalent vaccine but had completed a primary series of COVID-19 vaccination and had their last monovalent dose ≥60 days before 9/1/2022. Inclusion of these events helps to adjust for temporal trends (seasonality). The same NBR population was used in overall bivalent analyses as well as bivalent analyses stratified by co-administration of influenza vaccine. <sup>□</sup> Had SARS-CoV-2 infection (i.e., SARS-CoV-2 positive laboratory test or a COVID-19 diagnosis) during the year prior (08/31/2021–08/31/2022).

**eTable 3.** Numbers of Electronically Identified Ischemic Stroke Events and Relative Incidences in the 21 Days after Moderna Bivalent COVID-19 Vaccination.

|  | **All ages** |  |  |  | **<65 years old** |  |  |  | **≥65 years old** |  |  |  |
| --- | --- | --- | --- | --- | --- | --- | --- | --- | --- | --- | --- | --- |
|  | Number of events |  |  |  | Number of events |  |  |  | Number of events |  |  |  |
|  | Risk interval | Control interval | NBR<sup>§</sup> | Relative incidence (95% CI) | Risk interval | Control interval | NBR<sup>§</sup> | Relative incidence (95% CI) | Risk interval | Control interval | NBR<sup>§</sup> | Relative incidence (95% CI) |
| **Overall** | 82 | 745 | 3049 | 0.91 (0.71–1.15) | 13 | 179 | 1278 | 0.63 (0.35–1.14) | 69 | 566 | 1771 | 0.99 (0.76–1.29) |
| With history of SARS-CoV-2<sup>□</sup> | 10 | 129 | 565 | 0.60 (0.32–1.14) | 3 | 33 | 292 | 0.87 (0.25–2.97) | 7 | 96 | 273 | 0.52 (0.25–1.10) |
| Without history of SARS-CoV-2 | 72 | 616 | 2484 | 0.98 (0.75–1.28) | 10 | 146 | 986 | 0.58 (0.30–1.13) | 62 | 470 | 1498 | 1.11 (0.83–1.47) |
| **Co-administration of influenza vaccine, overall** | 8 | 67 | 3049 | 0.59 (0.27–1.28) | 4 | 19 | 1278 | 1.10 (0.37–3.21) | 4 | 48 | 1771 | 0.42 (0.14–1.27) |
| With history of SARS-CoV-2<sup>□</sup> | 1 | 16 | 565 | 0.34 (0.04–2.67) | 1 | 6 | 292 | 1.03 (0.12–8.79) | 0 | 10 | 273 | N/A |
| Without history of SARS-CoV-2 | 7 | 51 | 2484 | 0.65 (0.28–1.54) | 3 | 13 | 986 | 1.07 (0.31–3.63) | 4 | 38 | 1498 | 0.52 (0.16–1.67) |
| **No co-administration of influenza vaccine, overall** | 74 | 678 | 3049 | 0.95 (0.74–1.23) | 9 | 160 | 1278 | 0.53 (0.27–1.07) | 65 | 518 | 1771 | 1.07 (0.81–1.41) |
| With history of SARS-CoV-2<sup>□</sup> | 9 | 113 | 565 | 0.64 (0.33–1.25) | 2 | 27 | 292 | 0.79 (0.18–3.54) | 7 | 86 | 273 | 0.59 (0.28–1.24) |
| Without history of SARS-CoV-2 | 65 | 565 | 2484 | 1.03 (0.78–1.35) | 7 | 133 | 986 | 0.49 (0.22–1.07) | 58 | 432 | 1498 | 1.19 (0.89–1.60) |

<sup>§</sup>Non-bivalent recipients (NBR) were eligible individuals who did not receive a bivalent vaccine but had completed a primary series of COVID-19 vaccination and had their last monovalent dose ≥60 days before 9/1/2022. Inclusion of these events helps to adjust for temporal trends (seasonality). The same NBR population was used in overall bivalent analyses as well as bivalent analyses stratified by co-administration of influenza vaccine. <sup>□</sup> Had SARS-CoV-2 infection (i.e., SARS-CoV-2 positive laboratory test or a COVID-19 diagnosis) during the year prior (08/31/2021–08/31/2022).

